# Evaluation of plasma tau and neurofilament light chain biomarkers in a 12-year clinical cohort of human prion diseases

**DOI:** 10.1101/2020.07.27.20157594

**Authors:** Andrew GB Thompson, Prodromos Anastasiadis, Ronald Druyeh, Ines Whitworth, Annapurna Nayak, Akin Nihat, Tze How Mok, Peter Rudge, Jonathan DF Wadsworth, Jonathan Rohrer, Jonathan M Schott, Amanda Heslegrave, Henrik Zetterberg, John Collinge, Graham S Jackson, Simon Mead

## Abstract

Prion diseases are fatal neurodegenerative conditions with highly accurate CSF and imaging diagnostic markers, but major unmet needs for blood biomarkers. Using ultrasensitive immuno-assays, we measured tau and neurofilament light chain (NfL) protein concentrations in 709 plasma samples taken from 377 individuals with prion disease during a 12-year prospective clinical study, alongside healthy and neurological control groups. This provides an unprecedented opportunity to evaluate their potential as biomarkers. Plasma tau and NfL were increased across all prion disease types. For distinguishing sCJD from control groups including clinically-relevant “CJD mimics”, both show considerable diagnostic value. In sCJD, NfL was substantially elevated in every sample tested, including during early disease with minimal functional impairment and in all follow-up samples. Plasma tau was independently associated with rate of clinical progression in sCJD, while plasma NfL showed independent association with severity of functional impairment. In asymptomatic PRNP mutation carriers, plasma NfL was higher within 2 years of symptom onset than in samples taken earlier. We present biomarker trajectories for 9 individuals healthy at enrolment who developed symptoms during follow-up, showing potential for plasma NfL as a “proximity marker”. We conclude that plasma tau and NfL have potential to fill key unmet needs for biomarkers in prion disease: as an outcome for clinical trials (NfL and tau); for predicting onset in at-risk individuals (NfL); and as an accessible test for earlier identification of patients that may have CJD and require more definitive tests (NfL). Further studies should evaluate their performance directly in these specific roles.

## INTRODUCTION

The human prion diseases are a clinically and aetiologically heterogeneous group of neurodegenerative conditions, accounting for around 1 in 5000 deaths in the UK^1^. They share a core molecular pathology of templated conversion of the normal prion protein into abnormal disease-associated conformational states, but this process can be initiated in different ways^2^. In sporadic Creutzfeldt-Jakob disease (sCJD), the most common form, it appears to result from a random protein-misfolding event. In inherited prion diseases (IPD), it results from mutation of the prion protein gene (PRNP). In the acquired prion diseases, it results from exposure to exogenous prions either through diet (variant CJD (vCJD)) or through medical interventions (iatrogenic CJD (iCJD)).

Most patients with prion disease present with a strikingly rapid neurocognitive decline leading to a state of akinetic-mutism and death within a few months^3^. This rapid neurodegeneration results in dramatic increases in the concentrations of intra-neuronal proteins in the cerebrospinal fluid (CSF), which have historically provided valuable diagnostic biomarkers (e.g. 14-3-3 and tau proteins). However, in atypical sCJD cases, many forms of IPD, and in vCJD and iCJD, progression is typically slower, and the diagnostic value of these non-specific markers is weaker. More sophisticated diagnostic assays have now been developed (most notably the CSF RT-QuIC assay^4^), which when combined with improved brain imaging methods (particularly diffusion-weighted MRI) and PRNP gene sequencing allow the majority of patients with sCJD to be diagnosed with exceptional levels of accuracy^5^.

Nevertheless, a number of important roles for biomarkers in prion disease remain unfulfilled. Whilst small molecules that were promising in pre-clinical studies proved not to be effective in trials, immunotherapeutic and antisense oligonucleotide approaches to developing disease-modifying therapeutics for prion disease have shown substantial promise at pre-clinical stages^6-11^, and passive immunotherapy with a humanised anti-PrP^C^ monoclonal antibody is being clinically evaluated ^12^. However, there are a number of challenges in conducting clinical trials in prion disease, and overcoming these is vital alongside therapeutic development. The rarity and rapidity of sCJD often lead to the diagnosis being made at an advanced stage of disease, when substantial neurological damage has already occurred. A test using a readily accessible analyte applicable in the early stages could facilitate earlier diagnosis avoiding unnecessary investigation and hospital stay and enabling early enrolment to trials. The heterogeneity of prion diseases makes the use of clinical outcome measures in trials of feasible size challenging^13^, and a quantitative biomarker of disease activity that could act as a secondary outcome measure of biological treatment effect would be valuable. Identifying asymptomatic individuals carrying PRNP mutations presents the theoretical opportunity to give treatments to prevent disease. The wide and unexplained variation in age of symptom onset in all forms of IPD means that it is not currently feasible to use disease onset as an outcome14 and preventive treatments might need to be given for decades in at-risk individuals. A biomarker of proximity to disease onset might help overcome these problems.

Brain-derived markers of neurodegeneration measured in blood are potential candidates for these roles. Although they may lack diagnostic specificity, this does not prevent them from filling useful roles, alongside the existing specific diagnostics. Ultrasensitive immuno-assay methods, such as the Single molecule array platform (*Simoa*), now allow the accurate quantification of various brain-derived proteins in blood. Amongst the most promising of these are the tau (total-tau throughout) and neurofilament light chain (NfL) proteins15. Their concentrations in blood are elevated in a range of neurological conditions, including prion diseases16-18. In several contexts they provide quantitative measures of brain pathology: this includes providing a therapeutic biomarker for a central nervous system pathology (NfL in multiple sclerosis^19^), and a marker of pre-symptomatic neurodegenerative disease activity (NfL in carriers of mutations causing familial Alzheimer’s^20^ and Huntington’s diseases^21^).

Here we evaluate tau and NfL in the context of a large prospective natural history cohort of all types of human prion disease in the UK, providing an unprecedented opportunity to study their relationship to clinical progression, and to assess their potential to play key roles in future research aiming to establish effective treatments for these devastating diseases.

## METHODS

### Participants and plasma samples

Individuals were included if they were enrolled in the National Prion Monitoring Cohort study22 and/or the PRION-1 trial7 (collectively “the Cohort” hereafter), and had at least one suitable plasma sample available for testing (with appropriate research consent). These included patients with symptomatic prion disease (sCJD, IPD, iCJD and vCJD – diagnosed by standard criteria^23^), asymptomatic carriers of pathogenic *PRNP* mutations, and healthy control (HC) volunteers (friends or relatives of prion disease patients, excluding those at risk of carrying *PRNP* mutations).

In addition, several sets of neurological disease control samples were included. Samples from patients that were referred to the NHS National Prion Clinic on suspicion of having prion disease, but in whom an alternative diagnosis was made, were included to provide a clinically relevant diagnostic control group (“CJD mimics”). These included patients with a range of diagnoses, made on the basis of conclusive laboratory, neuropathology or clinical evidence (see Table 1 notes). All these patients were included in a recent clinical publication5. Samples from patients with AD and FTD, diagnosed according to contemporary diagnostic criteria at a specialist cognitive disorders clinic, were also included, to allow comparisons with other diagnostic groups where these biomarkers are known to be abnormal. Collection, transport, fractionation and storage of blood samples were carried out using standardised protocols. After fractionation, plasma samples were stored at −80°C until thawing immediately prior to use. These studies were approved by the local research ethics committee of UCL Institute of Neurology and the National Hospital for Neurology and Neurosurgery (NHNN).

**TABLE 1.**
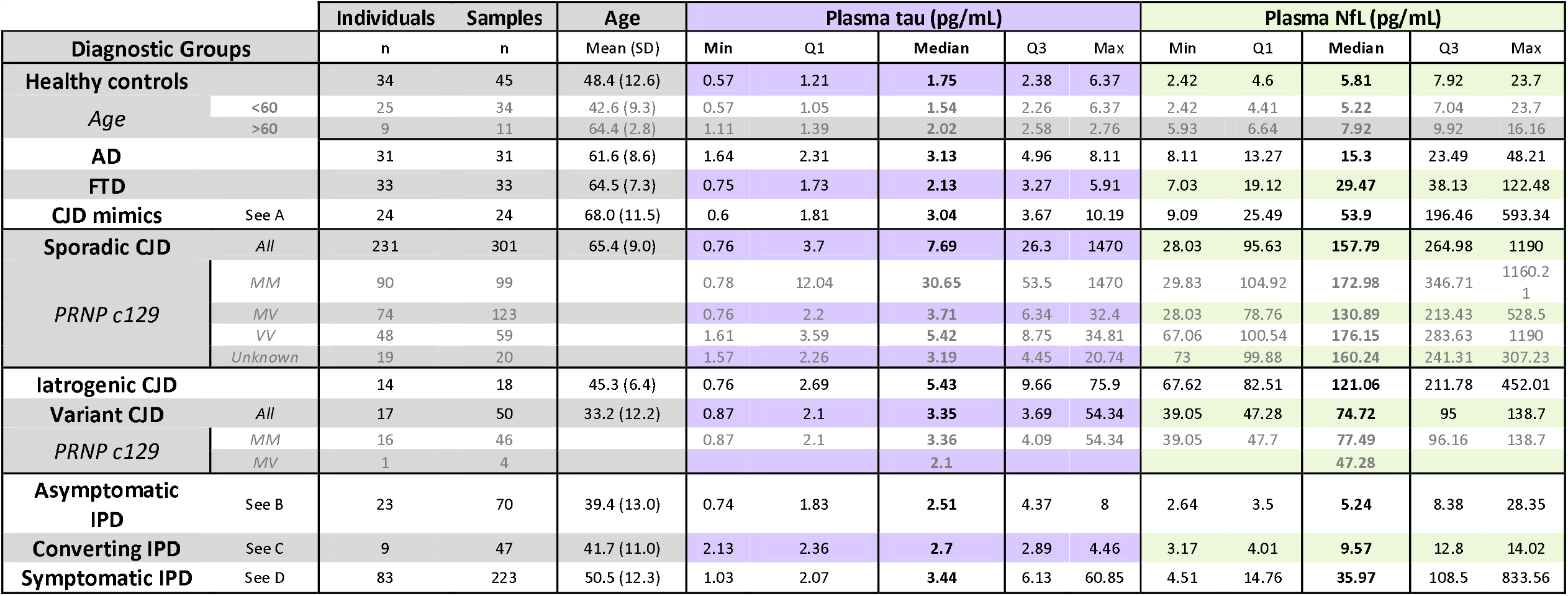
Summary of participants and plasma samples included in the study, and summary statistics for plasma tau and NfL concentrations for each diagnostic group. Biomarker summary statistics based on earliest available sample from all individuals. **A** – CJD mimic group includes patients with the following diagnoses (n): Lewy Body Dementia (8), Alzheimer’s disease (AD) (6), autoimmune encephalitis (6), AD and cerebrovascular (2), CNS lymphoma (2), cerebrovascular disease (1), frontotemporal dementia with motor neurone disease (1), progressive multifocal leukoencephalopathy (1), familial dementia with no identified cause (1), hepatic encephalopathy (1), unknown, not prion (1); **B** - P102L (9), E200K (7), D178N (2), Y163X (2), 5OPRI (1), 6OPRI (1), A117V (1); **C** - 5OPRI (1), 6OPRI (1), D178N (1), E200K (1), P102L (5); All individuals with “Converting IPD” were asymptomatic when the blood sample included in this dataset was taken, but subsequently developed symptomatic inherited prion disease during follow up (range 0 to 8 years from first sample); **D** - P102L (26), 6OPRI (19), E200K (10), 5OPRI (7), D178N (6), A117V (5), 4OPRI (3), Y163X (3), E196K (1), P105L (1), Q212P (1), V210I (1).

### Prospective clinical data

All patients with prion disease had systematic clinical scales data gathered at the time of enrolment to the Cohort, and then had further prospective face-to-face and telephone assessments, between 2-weekly and annually, determined by their expected rate of disease progression. Full details of the clinical studies, including follow-up schedules, have been published^7,22^.

The MRC Prion Disease Rating Scale (“MRC Scale” hereafter) is a bespoke rating scale for rapidly progressive prion disease, validated as a linear instrument to track clinical progression in individual patients. It runs from 20 (no significant functional impairment) to 0 (bedbound, mute, with minimal awareness of surroundings)^22^. MRC Scale data are available for all prion disease patients included in this study, providing a useful metric of functional impairment/disease progression, although it should be noted that formal statistical validation was restricted to those with rapidly progressive forms of prion disease.

We have developed a linear mixed modelling approach using MRC Scale data to quantify the rate of clinical disease progression in individual patients13, “MRC Slope”, expressed as percentage loss of function per day. This method cannot be applied to patients with severe functional impairment (MRC Scale < 5) at the time of enrolment.

### Quantification of tau and NfL in plasma samples

Tau and NfL concentrations were measured in plasma using the *Simoa HD-1 Analyser* and the “*tau-2*.*0*” and “*NF-light*” assay kits (*Quanterix*), according to manufacturer’s instructions and as described previously^18^. Samples from each diagnostic group were assorted across different loading plates and assay runs. Samples were tested in duplicate, by taking two aliquots from the same plate well. For quality control, data were reviewed after testing and samples without two valid results or with a coefficient of variation (CV) over 15% between duplicates were re-tested. All “*tau-2*.*0*” testing kits were from a single batch. “*NF-light*” kits were taken from 2 separate batches due to a manufacturer issue: a full calibration sample set tested for each batch showed no significant variation between them.

### Statistical analysis

As seen in our own previous work and elsewhere, neither tau nor NfL was normally distributed within any of the diagnostic groups tested (including healthy controls), with a positive skew. After log transformation (base 10), log-tau and log-NfL were plausibly normally distributed within diagnostic groups. Non-parametric methods were used for the primary analyses comparing groups, and log-transformed values were used for all other statistical analyses. Linear mixed effects modelling of changes in log-transformed biomarker concentrations as the disease progresses was done in two ways: once using change over time (days from death), and once using change with MRC Scale. We included fixed effects for age and *PRNP* codon 129 genotype (known phenotype modifiers in other contexts), and allowed random effects for slope and intercept to account for variation between individuals. Details of the models are given in the relevant Results sections below. Regression analysis of baseline log-transformed biomarker concentrations included both MRC Scale at the time of blood sampling and the individual’s MRC Slope as covariates, along with age, gender and *PRNP* codon 129 genotype. Analyses were carried out using *Microsoft Excel* and *Stata 12*.*1*.

## RESULTS

### Comparison of plasma tau and NfL between symptomatic prion disease and control groups

Patients and plasma samples included in the study are summarised in Table 1, Figure 1 and ROC curves (Figure 2). For both tau and NfL there were highly significant differences across diagnostic groups including HCs (Kruskal-Wallis, p=0.0001). Compared with the HC group, plasma tau concentrations were elevated in sCJD (Dunn’s *post hoc* pairwise comparison test with Bonferroni correction, z=8.7, p<0.0001), symptomatic IPD (z=4.5, p=0.0001) and iatrogenic CJD (z=3.8, p=0.0018), but not for vCJD (z=2.5, p=0.1669). Plasma tau in sCJD was higher than in AD (z=4.1, p=0.0005) and FTD (z=6.6, p<0.0001). No other prion disease groups were significantly different from either AD or FTD (all p>0.1). Tau was higher in sCJD than in symptomatic IPD (z=5.28, p<0.0001) and vCJD (z=3.4, p=0.0093), but no other pairwise comparisons amongst prion disease groups were significant (all p>0.5).

**Figure 1.**
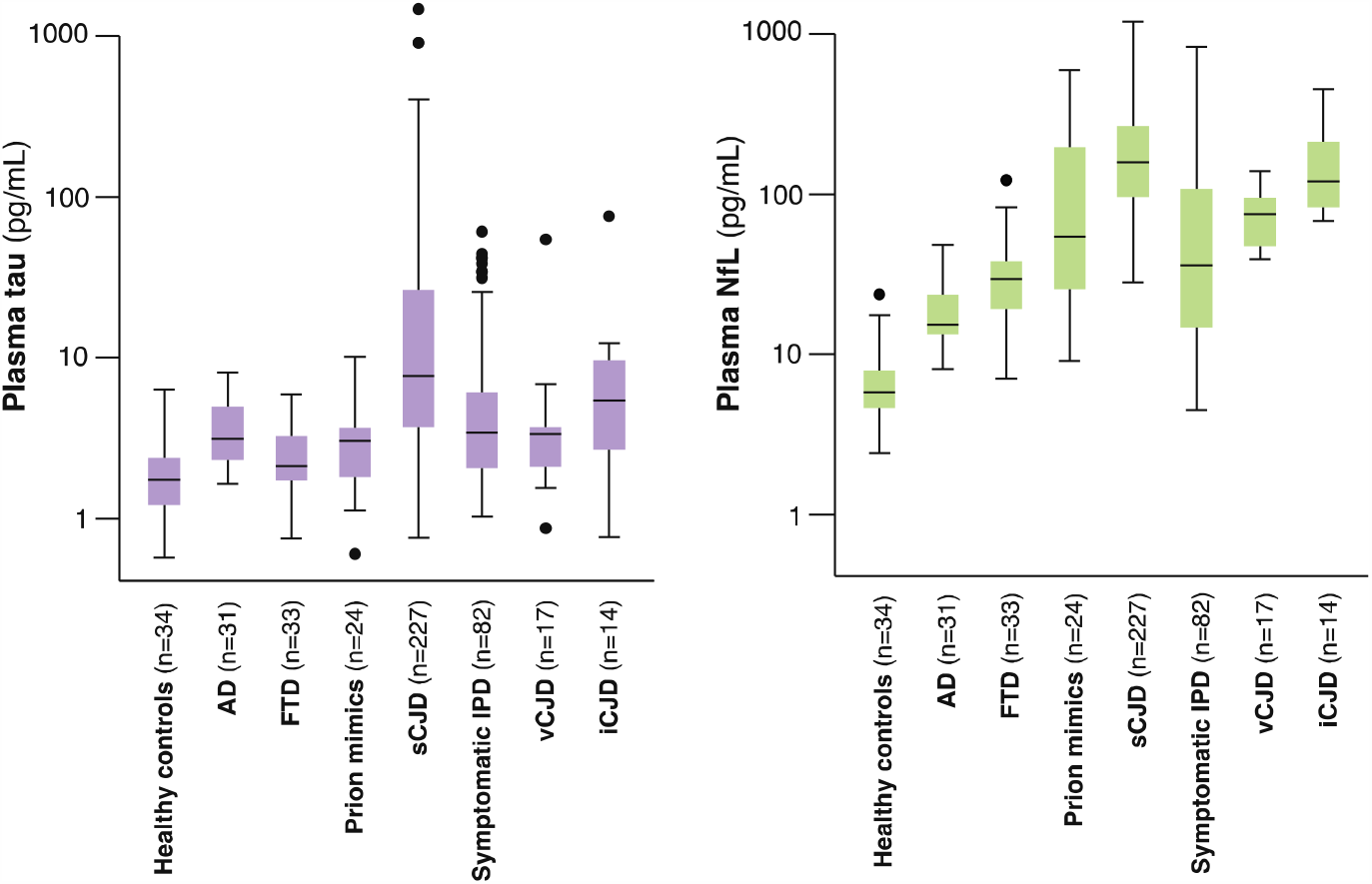
Plasma tau and NfL concentrations (shown on log scales) across symptomatic prion disease and control groups. Boxplots show median, upper and lower quartiles, and minimum and maximum excluding any outliers (defined using Tukey method, as more than 1.5 × Interquartile Range outside the quartile). Alzheimer’s disease (AD), frontotemporal dementia (FTD), inherited prion disease (IPD), sporadic, variant and iatrogenic Creutzfeldt-Jakob disease (sCJD, vCJD and iCJD).

**Figure 2.**
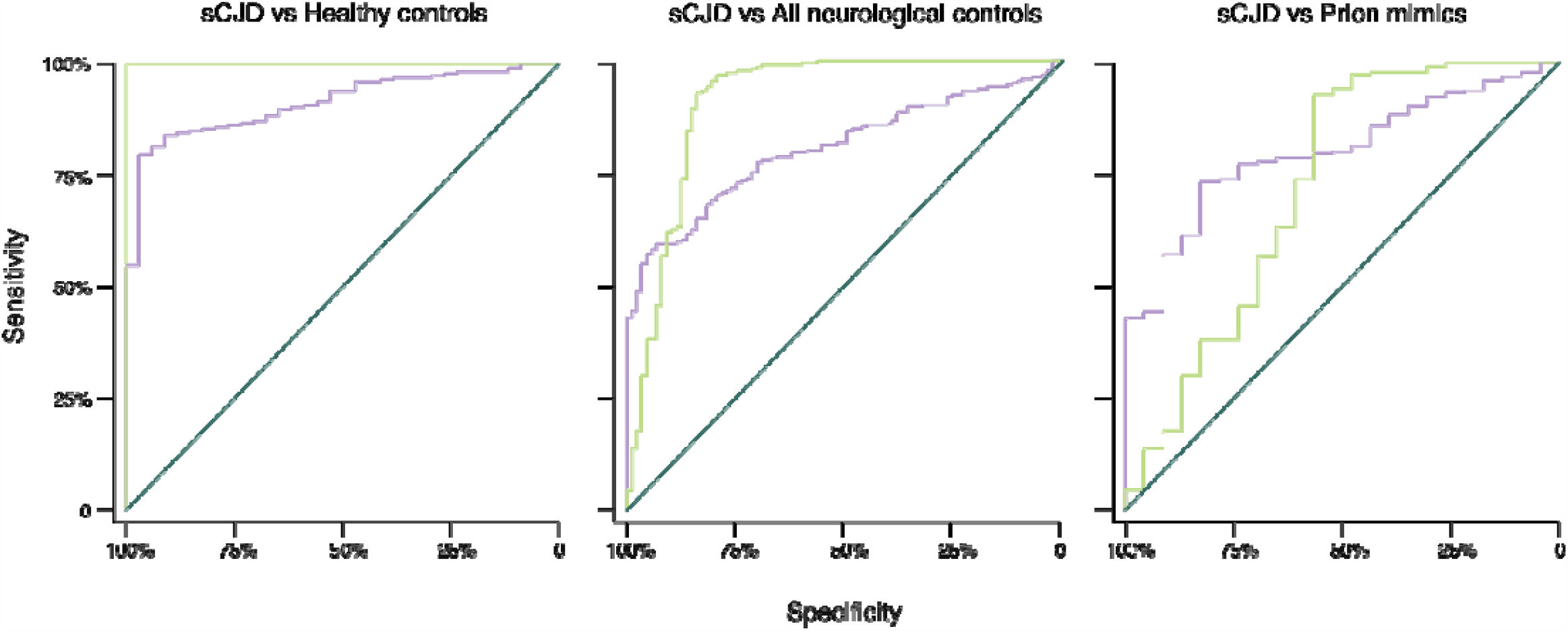
Receiver Operating Characteristic (ROC) curves. Plasma tau (purple) and NfL (green), for distinguishing sCJD from **(A)** Healthy controls, **(B)** all neurological disease controls (AD, FTD and prion mimics), and **(C)** prion mimics only. Areas under curves are given in Table 2 in the main paper.

Compared with the HC group, plasma NfL was elevated in sCJD (z=11.8, p<0.0001), symptomatic IPD (z=5.7, p<0.0001), vCJD (z=4.5, p=0.0001), and iatrogenic CJD (z=6.3, p<0.0001). Compared with the AD group, plasma NfL was higher in all prion disease types: sCJD (z=9.1, p <0.0001), symptomatic IPD (z=3.5, p=0.0062), vCJD (z=3.0, p=0.037) and iatrogenic CJD (z=4.9, p<0.0001). Comparing with the FTD group, it was higher in sCJD (z=7.7, p<0.0001) and iatrogenic CJD (z=3.9, p=0.0012), but there was no significant difference with symptomatic IPD or vCJD (both p>0.5). Amongst prion disease groups, NfL was higher in sCJD than in both symptomatic IPD (z=7.8, p<0.0001) and vCJD (z=3.28, p=0.014), but no other pairwise comparisons showed significant differences (all p>0.05).

The ages of HCs in our study span the wide range seen in the different prion disease groups, but do not match each individual group perfectly. It is important to consider that biomarkers vary with age in HC: plasma NfL tends to increase by about 2.2% per year in HCs^15^. Table 1 includes separate biomarker distributions for younger and older HCs, showing the relatively modest increase in the older subset.

We carried out Receiver Operating Characteristic (ROC) analyses, and calculated diagnostic parameters using optimal cut-off values derived from the ROC analysis (those maximising the Youden index^24^)(Table 2 and Figure 2). NfL separated sCJD cases from HCs completely and performed well in distinguishing sCJD from the mixed neurological disease control group (ROC AUC=0.91). However, tau appeared most effective for the more clinically relevant role of distinguishing sCJD from the CJD mimic group (ROC AUC=0.81).

**Table 2.** Diagnostic parameters for plasma tau and NfL in sCJD, derived from Receiver Operating Characteristic (ROC) analysis.

| **sCJD...** | **vs Healthy controls** |  | **vs All non-prion disease** |  | **vs 'CJD mimics'** |  |
| --- | --- | --- | --- | --- | --- | --- |
| **Plasma biomarker** | **tau** | **NfL** | **tau** | **NfL** | **tau** | **NfL** |
| **Area under ROC curve** | **0.913** | **1.0** | **0.808** | **0.912** | **0.809** | **0.724** |
| **Cut-off (pg/mL)**<br>(Youden method) | **>3.27** | **>25.87** | **>6.11** | **>60.65** | **>3.81** | **>60.65** |
| **Sensitivity**<br>(95% C.I.) | **80%**<br>(73% - 87%) | **100%**<br>(n/a) | **57%**<br>(42% - 73%) | **93%**<br>(89% - 97%) | **74%**<br>(59% - 88%) | **93%**<br>(88% - 98%) |
| **Specificity**<br>(95% C.I.) | **97%**<br>(90%-100%) | **100%**<br>(n/a) | **95%**<br>(82% - 100%) | **84%**<br>(75% - 93%) | **83%**<br>(68% - 99%) | **57%**<br>(39% - 74%) |

### Plasma tau and NfL as markers of disease progression in sCJD

We explored the relationship between biomarker concentrations and disease progression in sCJD by crosssectional and longitudinal analysis. For the cross-sectional analysis, data were plotted against time (days to death) and MRC Scale (Figure 3) using the first sample available. In the baseline analysis of log-tau, higher concentrations were found in patients with more rapid decline (R^2^=0.36; MRC Slope predictor, z=7.15, p<0.001), while stage of progression (MRC Scale predictor) showed only weak evidence (z=-2.00, p=0.048). There was no independent effect of PRNP codon 129 genotype, suggesting that the difference in plasma tau between genotypes (see Table 1) results from their different rates of progression rather than being directly related to prion strain type. In a subset of 54 patients with molecular strain typing (PrP^Sc^ typing) available^25^, we found that MM-2 patients tended to have higher plasma tau concentration than MM-3 (London classification^26^; equivalent to types 1 and 2 by Parchi classification^27^), again in keeping with their rates of progression (see Supplementary Table).

**Figure 3A.**
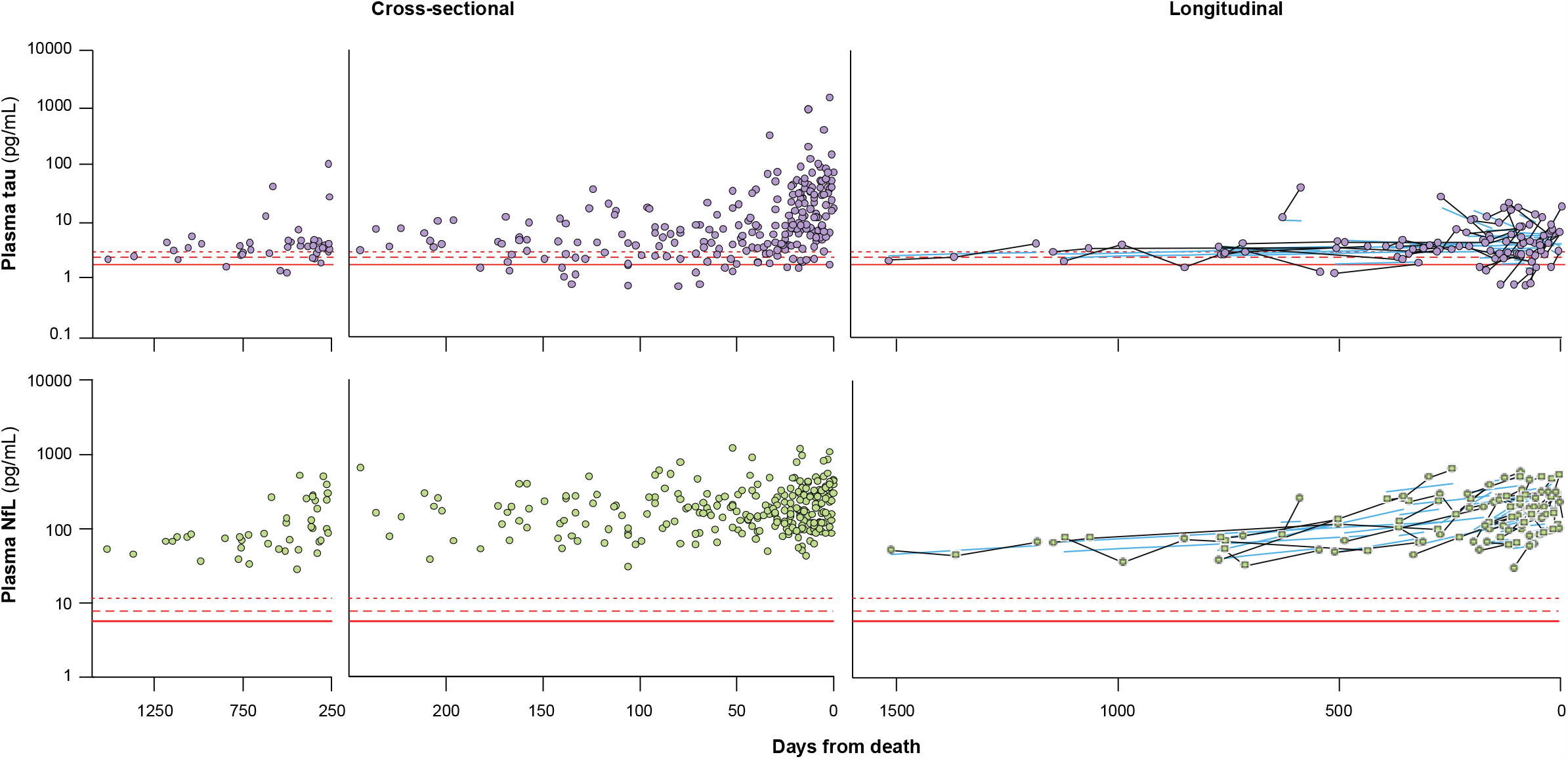
Relationship of plasma tau and NfL to number of days from death in sporadic CJD, in cross-sectional dataset (earliest available sample from each patient – left panels) and longitudinal dataset (all samples from patients with more than one available – right panels). Red continuous, dashed and dotted lines show 50th, 75th and 90th centiles of healthy controls respectively. Blue lines on the longitudinal charts show the linear mixed effects model fits for each individual (see text). Patients with very rapidly progressive disease, typically enrolled to the Cohort at a late stage of disease, are less likely to have more than one blood sample available, so are under-represented in the longitudinal sample set.

**Figure 3B.**
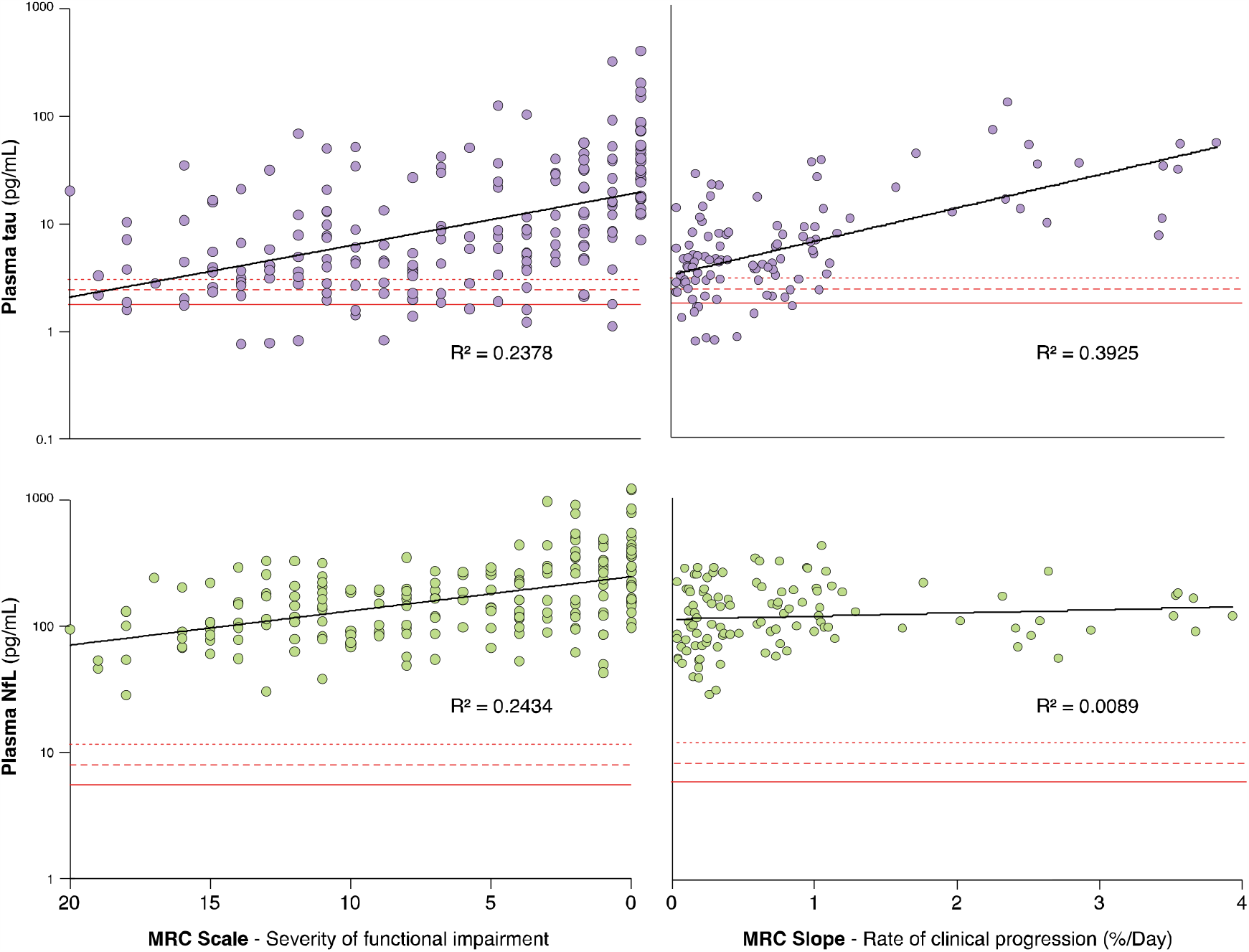
Relationship of plasma tau and NfL to measures of disease progression in sporadic CJD. Left panels show biomarkers (log scale) plotted against MRC Scale, which measures severity of functional impairment. Right panels show biomarkers (log scale) plotted against MRC Slope, which provides a measure of the rate of clinical progression. The earliest sample from each patient with necessary data available is included. As MRC Slope cannot be modelled for patients with a baseline MRC Scale < 5, fewer patients are represented in MRC Slope charts, and the missing cases are skewed towards those enrolled at an advanced stage of disease, and those with very rapid clinical progression.

In contrast, for log-NfL, stage of progression was a strong predictor (R2=0.36; MRC Scale predictor, z=-5.82, p<0.001) with higher NfL concentrations at more advanced disease stages, while there was no compelling evidence for an effect of MRC Slope (z=1.78, p=0.077, Figure 3). As expected, age also had an independent effect on log plasma NfL (for details see Supplementary Table 1).

In longitudinal analysis, plasma NfL was substantially raised throughout the follow-up period in all patients, with no examples of NfL dropping into the range of concentrations seen in HCs (Figure 3, see Supplementary Figure 1 for variation in longitudinal sample pairs). Here, we again found a significant independent effect of stage of progression (days from death (p<0.001), MRC Scale (p<0.001)) on log plasma NfL. There was also some variation between codon 129 groups, with more rapid increase over time in VV than MV patients (p=0.008). The individual random effects had a proportionally greater impact on intercept than on slope, suggesting that while some individuals have higher NfL at equivalent stages of disease, the rate of change is relatively consistent between individuals within codon 129 genetic groups, and adjusted for age. In the models for tau, there was no independent effect of days from death, and the effect of MRC Scale was not compelling, considering multiple testing (p=0.016, Supplementary Table 2).

We explored whether baseline assessment of plasma tau or NfL concentration could help to predict subsequent rate of clinical disease progression in sCJD patients, and thereby have potential value for stratifying patients at enrolment to clinical trials. Adding baseline tau to existing models including baseline MRC Scale and *PRNP* codon 129 (as previously published^13^) might lead to some improvement in predicting rate of progression, but such an effect was probably driven by a specific subset of very rapidly progressive patients (those with MM codon 129 genotype), and the improvement was modest (Supplementary data).

### Plasma tau and NfL in inherited and acquired prion diseases

The study included 340 plasma samples from 115 *PRNP* mutation carriers, with a range of different mutations (see Table 1 notes). 9 individuals who were asymptomatic at the time of their earliest blood sample went on to develop symptoms of IPD during follow up, and for 6 of these individuals, samples from before and after symptom onset were tested in this study. The date of symptom onset was taken as the date at which individuals reported first experiencing any symptoms consistent with IPD, unless these subsequently resolved or proved to be due to an alternative cause.

To explore changes in the blood biomarkers as individuals approach the onset of neuropsychiatric symptoms of IPD, we grouped plasma samples as follows: (1) samples taken more than 2 years prior to symptom onset (including patients with a known date of onset, and those that are known to have remained asymptomatic for at least 2 years after the date of sampling): *74 samples from 23 individuals*; (2) samples taken within 2 years prior to symptom onset: *11 samples from 7 individuals*; (3) samples taken in early symptomatic IPD: after onset of symptoms, but while day-to-day functioning remained unimpaired: *22 samples from 13 individuals*; (4) samples taken after the development of IPD-related functional impairment: *220 samples from 83 individuals* (Figure 4).

**Figure 4A.**
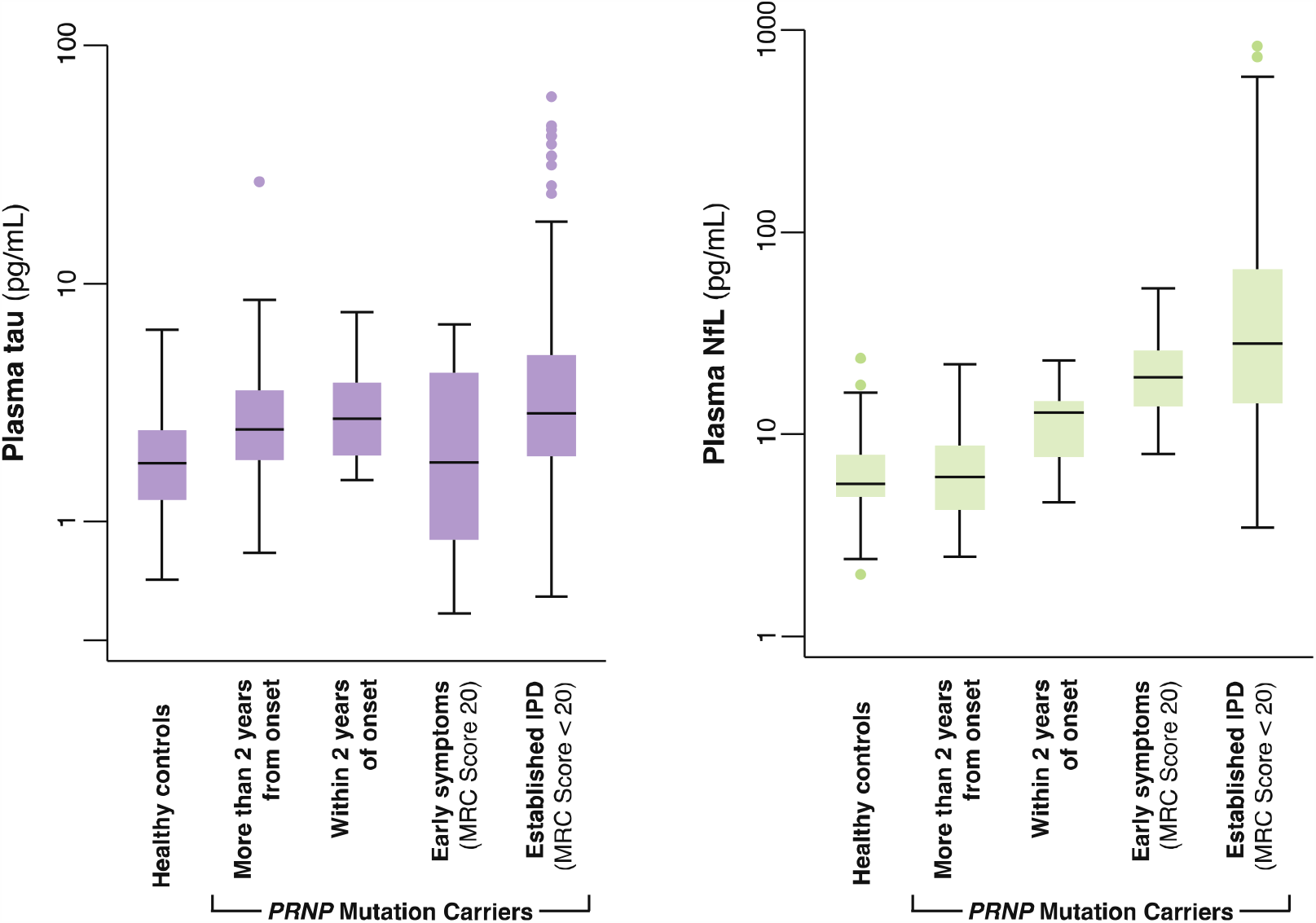
Plasma tau and NfL (log scales) at different stages in *PRNP* mutation carriers. NB - Multiple samples from some individuals are included within and between different groups in these charts, and proportions of different *PRNP* mutations varies between groups. Boxplots show median, upper and lower quartiles, and minimum and maximum excluding any outliers (defined using Tukey method, as more than 1.5 × Interquartile Range outside the quartile).

**Figure 4B.**
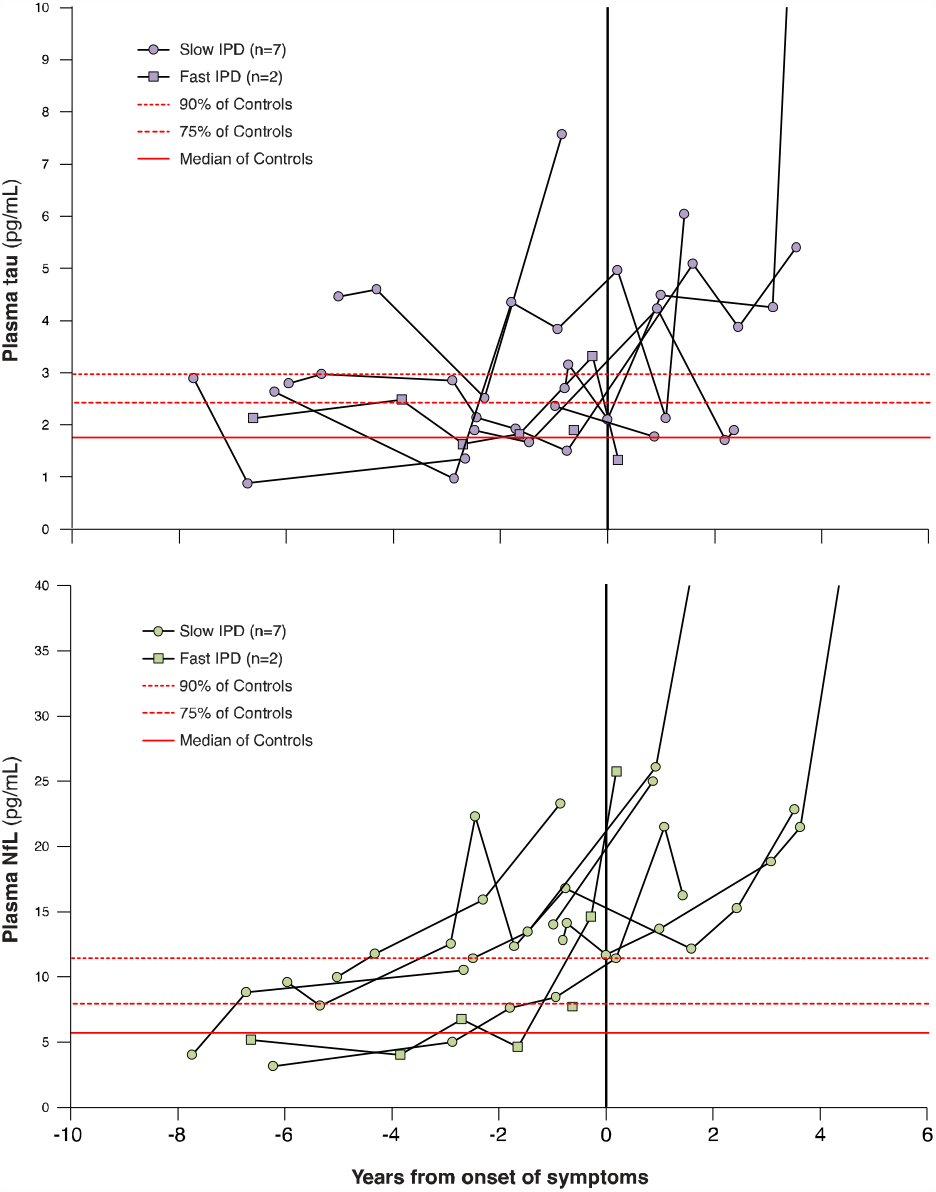
Individual plasma tau and NfL trajectories for *PRNP* mutation carriers that were asymptomatic at the time of their first sample, and subsequently developed symptoms of IPD during follow-up in the Cohort study. “Slow IPD”:P102L × 5, 5OPRI × 1, 6OPRI × 1. “Fast IPD”:E200K × 1, D178N × 1.

Plasma tau was modestly increased relative to HCs in all IPD sample groups, including those taken more than 2 years from onset (Mann-Whitney: *z*=-3.4,p=0.0007), but did not differ between groups (1), (2) and (3) above (Kruskal-Wallis: χ^2^=0.363,p=0.547). Plasma NfL concentrations were not different in samples taken more than 2 years from onset compared with HC samples (M-W: *z*=-0.222,p=0.824), but increased progressively between groups (1), (2) and (3) above (K-W: χ^2^=46.56,p=0.0001). Of particular note, NfL was higher in samples taken within 2 years of symptom onset than those taken more than 2 years from symptom onset (M-W: *z*=10.29, p=0.0013). The individual trajectories of plasma tau and NfL for those individuals with samples available prior to a known date of symptom onset are shown in Figure 4B, consistent with a predictive value of both the absolute NfL level, and rate of biomarker change.

In symptomatic IPD, *PRNP* mutations typically causing a rapid CJD-like disease phenotype (E200K, 4OPRI, E196K, V210I) were associated with very high concentrations of both tau and NfL in plasma, similar to those seen in sCJD, while the D178N mutation (typically causing either a CJD-like or fatal familial insomnia phenotype) gave slightly lower concentrations^28^. Mutations typically causing Gerstmann-Sträussler-Scheinker syndrome (P102L, A117V, P105L) seemed to result in a modest increase in tau and a relatively marked (though more variable) increase in NfL. Mutations typically causing a relatively slow cognitive decline (5OPRI, 6OPRI) produced more modest elevations. Changes in plasma biomarkers with disease progression in IPD were also studied (see Supplementary data).

Plasma tau and NfL concentrations for patients with vCJD and iCJD are plotted against time for both cross-sectional and longitudinal datasets (see Table 1, Figure 1, and Supplementary data). Briefly, biomarker concentrations were raised in iCJD and vCJD, but to a lesser extent than in sCJD, in line with their slower rates of progression. Longitudinally, NfL tended to remain elevated, as seen in sCJD.

## DISCUSSION

Studying plasma tau and NfL in a large cohort with rich, contemporaneous, prospective clinical data has provided a unique opportunity to evaluate their potential as biomarkers in human prion diseases. In sCJD, plasma NfL was markedly elevated in every sample tested, including those taken at early disease stages with minimal functional impairment, with no value overlapping HCs in the longitudinal study. Plasma tau and NfL had complementary associations with rate and stage of clinical progression. In a mixed cohort of asymptomatic *PRNP* mutation carriers, plasma NfL showed promising evidence of functioning as a proximity biomarker within two years of observed clinical onset. These observations suggest several future uses of blood biomarkers in human prion disease.

There have been major advances in laboratory diagnosis of prion disease in the last few years, driven primarily by the real-time quaking induced conversion (RT-QuIC) assay for CSF as a test for sCJD^29^. This is based on a highly disease-specific molecular process (seeded prion protein misfolding), and as a result confers substantial advantages over older CSF tests using surrogate markers of rapid neurodegeneration, particularly in terms of specificity. CSF analysis will remain vital in patients with suspected sCJD to look for evidence of other conditions for which specific treatments are available, such as autoimmune or viral encephalitis, and RT-QuIC provides exquisite specificity for CJD. We have shown that plasma NfL is substantially elevated even in sCJD patients with minimal functional impairment and in the early stages of atypically long disease courses. Plasma NfL may therefore have value as a triage test: flagging the possibility of sCJD or a CJD mimic in patients with cognitive disorders when found at high levels atypical for common dementias (e.g. >100pg/ml), and prompting consideration of specific CSF tests. Any approach of this sort would need to be studied prospectively and validated in the clinical setting in which it would be applied, with expanded comparison groups.

Plasma NfL is very substantially increased in all tested patients with sCJD, and importantly we have shown that even in longitudinal sample time courses across the spectrum of phenotypic heterogeneity, it was elevated in every sample of over 300 tested. This knowledge of the biomarker’s natural history is valuable when considering its potential as an outcome measure in therapeutic trials. Tau correlated with rate of clinical progression, although this appears to be driven mainly by very high values in extremely rapidly progressing patients and it added little to a predictive model that includes age, sex and *PRNP* codon 129. The concentrations of tau and NfL in blood will be influenced not only by their release from degenerating neurons, but also by their movement between different physiological compartments (brain extracellular fluid, CSF, blood) and their clearance from blood. If an experimental therapeutic agent affected these other processes, it may change biomarker levels independently of any effect on neurodegeneration. Plasma tau and NfL have some appealing and complementary properties as therapeutic markers, and notably different half-lives in blood^30^, but their suitability should be evaluated directly in patients receiving putative treatments. It would also be useful to study them in animal models in which successful treatment of prion disease can be achieved^11^.

As a result of the large and unexplained variation in the age of symptom onset in *PRNP* mutation carriers^14^, trial designs using timing of symptom onset as an outcome measure are challenging, as they require larger numbers of participants and a longer duration than might reasonably be achieved. For the same reason, we cannot use estimates of age at onset to measure biomarker performance. A “prodromal” or “proximity” biomarker could improve prospects for preventive trials because enrolled patients would be more likely to develop symptoms in the timeframe of a trial, and greater risks of treatment might be justified than in those without evidence of an active disease process. Plasma NfL was elevated in a group of IPD within two years of clinical onset and could fulfil this role. We have had the remarkable opportunity to study blood biomarkers longitudinally in individuals as they develop symptoms of prion disease over 12 years. Despite the size and duration of our Cohort Study, it included only a small number of patients in whom symptom onset was observed during follow up. It is also important to note that this includes individuals with several different *PRNP* mutations, which are associated with heterogeneous clinical phenotypes in their symptomatic phases. It is reasonable to speculate that different mutations might associate with different prodromal biomarker profiles. Differences in patient populations and length of follow up might explain differences between Cohorts^31,32^.

In summary, we report the performance of plasma NfL and tau biomarkers in a large natural history study of all forms of human prion disease. Future work might focus on specific functions and comparisons with other biomarkers, in appropriate settings, such as (1) the use of high levels of plasma NfL as a triage blood test in patients with cognitive disorders to prompt urgent referral to specialist care, (2) the use of both biomarkers in a clinical trial setting as secondary outcomes, and (3) further prospective study of NfL in longitudinal cohorts at-risk of inherited prion disease, particularly focussing on differences between different *PRNP* mutations, defining a prodromal window size and testing the value of biomarker trajectory rather than absolute level in prediction.

## Data Availability

All Nfl and t-tau biomarker data is available

## Acknowledgements

We would like to thank study participants and their relatives and carers for their contribution to Cohort studies. Richard Newton assisted with figure design. JC and SM are NIHR Senior Investigators. HZ is a Wallenberg Scholar. Professor Sarah Walker provided statistical advice.

